# In vivo myelin imaging and tissue microstructure in white matter hyperintensities and perilesional white matter

**DOI:** 10.1101/2021.11.23.21266731

**Authors:** Jennifer K. Ferris, Brian Greeley, Irene M. Vavasour, Sarah N. Kraeutner, Shie Rinat, Joel Ramirez, Sandra E. Black, Lara A. Boyd

## Abstract

**Introduction:** White Matter hyperintensities (WMHs) relate to cognitive decline in aging, due to their deleterious effect on white matter structure. Diffusion tensor imaging (DTI) detects changes to white matter microstructure, both within the WMH and extending in a penumbra-like pattern in surrounding (perilesional) normal appearing white matter (NAWM). However, DTI markers are not specific to tissue components, complicating interpretation of previous DTI findings. Myelin water imaging is a novel imaging technique that provides specific markers of myelin content (myelin water fraction: MWF) and interstitial fluid (geometric mean T2: GMT2). Here we combined DTI and myelin water imaging to examine tissue characteristics in WMHs and perilesional NAWM. Our goal was to establish a multimodal neuroimaging approach to track WMH progression.

**Methods:** 80 individuals (47 older adults and 33 individuals with chronic stroke) underwent neuroimaging and tissue segmentation. To measure perilesional NAWM, WMH masks were dilated in 2mm segments up to 10mm in distance from the WMH to measure perilesional NAWM. Fractional anisotropy (FA), mean diffusivity (MD), MWF, and GMT2 were extracted from WMHs and perilesional NAWM. We examined whether white matter metrics showed a spatial gradient of effects in perilesional NAWM, as a function of Distance from the WMH, and Group. We tested whether white matter metrics in the WMH lesion related to severity of cerebrovascular disease across the whole sample, indexed by whole brain WMH volume.

**Results:** We observed a spatial gradient of higher MD and GMT2, and lower FA, in perilesional NAWM and the WMH. In the chronic stroke group, MWF was reduced in the WMH lesion but did not show a spatial gradient in perilesional NAWM. Across the whole sample, white matter metrics within the WMH lesion related to whole-brain WMH volume; MD and GMT2 increased, and MWF decreased, with increasing WMH volume.

**Conclusions:** NAWM adjacent to WMHs exhibits characteristics of a transitional stage between healthy NAWM tissue and WMH lesions. This was observed in markers sensitive to interstitial fluid, but not in the specific marker of myelin concentration (MWF). Within the WMH, interstitial fluid was higher and myelin concentration was lower in individuals with more severe cerebrovascular disease. Our data suggests that fluid-sensitive imaging metrics (such as DTI) can identify NAWM at high risk of conversion to a WMH. In contrast, specific markers of myelin concentration (e.g., MWF) can be used to measure levels of demyelination in the WMH itself, which may be a useful marker for disease staging of advanced WMHs.

## Introduction

White Matter Hyperintensities (WMHs) are the most frequent incidental MRI finding in aging^1^. WMHs are present in >70% of individuals above the age of 60^1^, and high WMH volumes relate to cognitive decline^2^, a doubled risk of dementia^3^, and a tripled risk of stroke^3^. The identification of individuals at high risk of WMH accumulation will help target disease prevention efforts, but the early identification of WMHs is challenging. The cognitive impacts of WMHs are typically subtle or silent until late disease stages^4^, meaning there is a unique role for neuroimaging in the early identification of WMHs^5^. WMHs are histopathologically heterogeneous^6^, thus the ideal quantitative neuroimaging markers will be sensitive to disease progression and reflect disease pathology. Diffusion Tensor imaging (DTI) has been suggested as a potential sensitive marker of preclinical WMHs^5,7^, but DTI is limited by its lack of specificity to tissue pathology. Here we combine DTI with myelin water imaging, a more specific imaging marker of white matter characteristics, to investigate changes to white matter characteristics across a spectrum of cerebrovascular disease. We analyzed data from the WMH itself, and in surrounding normal appearing white matter (NAWM), tissue that is at high risk of future conversion to a WMH.

Histopathological studies indicate that WMH pathology evolves over the course of the disease^8,9^. One theory of WMH development suggests that early in disease pathology, venous collagenosis^10^ and increased blood brain permeability^11^ cause increased local interstitial fluid. This increased interstitial fluid then triggers a pathological cascade to glial (including oligodendroglial) and neuronal cells leading to demyelination and axonal damage^12,13^. Early interstitial fluid accumulation is potentially reversible, but usually the trend is for progression towards irreversible myelin loss and neuronal damage^8^. Importantly, both increased interstitial fluid and changes to tissue structure, such as demyelination and axonal loss, will appear bright (hyperintense) on a T2-weighted scan. This means that T2-weighted imaging alone can provide one measure of the *severity* of WMHs (i.e., their volume), but cannot quantify the *stage* of disease (i.e., the underlying shift from fluid accumulation to demyelination).

DTI can reveal subtle changes to white matter related to WMH-pathology. When comparing WMHs to NAWM, in the WMH fractional anisotropy (FA; a measure of directionality of diffusivity in the voxel) is lower and mean diffusivity (MD; a measure of mean diffusivity across all 3 principal directions in the voxel) than in NAWM (for review see Pasi, *et al*.^14^). Further, WMH-related microstructural changes extend beyond the WMH lesion itself into adjacent NAWM^15,16^; NAWM immediately bordering the WMH shows WMH-like characteristics that gradually level off with increasing distance from the WMH. This non-linear spatial effect in DTI metrics has been termed a “WMH penumbra”^15^ and suggests that DTI can detect early microstructural changes in NAWM that are subvisible on a T2-weighted scan. However, while changes to DTI microstructure are often interpreted as reflecting demyelination, FA and MD are also impacted by multiple factors beyond damage to myelinated axons, such as underlying fiber architecture, the presence of glial cells, and changes in local fluid^17,18^. This is an important caveat in interpreting DTI research of WMHs, where both increased interstitial fluid and demyelination are implicated in disease pathogenesis. Duering *et al*.^19^ found that the component of the DTI signal that reflects isotropic water (free water) explained the altered FA and MD within WMHs. However, the free water signal employed by Duering *et al*.^19^ was derived from DTI metrics and therefore does not give a specific measure of myelination to differentiate interstitial fluid from demyelination changes in WMHs and surrounding tissue.

Myelin water imaging is a novel imaging technique that measures the T2 distributions of the magnetic resonance signal^20^, and is an ideal imaging technique to examine WMHs and surrounding NAWM. Unlike DTI, myelin water imaging is specific to tissue characteristics. Myelin water imaging is based on differentiating two primary signals from the T2 distribution. The first signal is a short T2 component (between 10 to 40ms), from water trapped in the myelin bilayers wrapping the axon. The area in the short T2 distribution is normalized by the area of total T2 distribution to give myelin water fraction (MWF). MWF has been histologically validated at 1.5T^21^ and 7T^22^ as a sensitive and specific marker of myelin concentration. The second signal is a long T2 component (between 40 to 200ms) from intracellular and extracellular water, calculated as the geometric mean T2 (GMT2; amplitude-weighted mean on a logarithmic scale^23^). Myelin water imaging can therefore confirm whether the “WMH penumbra” of altered FA and MD in WMHs and surrounding NAWM is more likely to reflect demyelination (assessed by MWF) or changes to interstitial fluid (assessed by GMT2). Myelin water imaging may also be able to stage disease severity within the WMH itself, giving a more sensitive metric of WMH disease progression beyond simple WMH volume.

Our goal was to establish a multimodal neuroimaging approach to give insight into WMH disease staging and progression. To accomplish this, we combined DTI and myelin water imaging to examine subvisible tissue changes in WMHs and surrounding NAWM (hereafter referred to as “perilesional NAWM”). Our sample represented a spectrum of cerebrovascular disease severity; we included a cohort of older adults with mild WMHs, and a cohort of individuals with chronic stroke with more severe WMHs. Our primary aim was to test if WMHs impact DTI and myelin water imaging metrics, both in the WMH itself and in perilesional NAWM. We hypothesized we would observe FA increasing and MD decreasing quadratically in NAWM as a function of distance from the WMH. We further hypothesized we would see GMT2 decreasing quadratically as a function of distance from the WMH, indicating elevated interstitial fluid. We predicted the effect in MWF would vary depending on the severity of cerebrovascular disease. Specifically, we hypothesized that: there would be no spatial gradient of MWF early in cerebrovascular disease (i.e., older adults), whereas individuals with advanced cerebrovascular disease (i.e., individual with chronic stroke) would show a spatial gradient of MWF, indicating myelin loss. Our secondary aim was to test whether DTI and myelin water imaging metrics in the WMH related to the severity of WMH disease, assessed by whole brain WMH volume. We hypothesized that white matter metrics in the WMH would relate to whole-brain WMH volume, indicating that pathology in the WMH evolves with increasing severity of cerebrovascular disease

## Materials and methods

### Participants

Data for this study were pooled from two previous research studies as a secondary analysis. We included 50 otherwise healthy older adults and 33 individuals with chronic stroke who received multimodal neuroimaging between 2016 and 2020. Participants were initially recruited through community postings in Vancouver and the Greater Vancouver Area. All participants were between the ages of 40-80 years. Participants were excluded if they: 1) had a history of seizure/epilepsy, head trauma, a major psychiatric diagnosis, neurodegenerative disorders, or substance abuse and/or 2) reported any contraindications to MRI. Older adults were excluded if they had extensive cerebral small vessel disease defined by WMH volumes ≥10 mL, which has been posited as a minimum clinically-significant threshold for WMH-related cognitive and neurological changes^24,25^. Individuals with chronic stroke were excluded if they were ≤6 months post a clinically diagnosed stroke (i.e., participants were included in the chronic phase of recovery). Montreal Cognitive Assessment (MoCA) test scores were acquired on all study participants^26^. Informed consent was obtained for each participant in accordance with the Declaration of Helsinki. The University of British Columbia research ethics boards approved all aspects of study protocols.

### MRI acquisition

MRI images were acquired on 3.0T Phillips Achieva or Elition scanners (Philips Healthcare, Best, The Netherlands), at the University of British Columbia MRI Research Centre with an eight-channel or thirty-two-channel sensitivity encoding head coil, respectively, and parallel imaging. The following structural imaging sequences were acquired: a 3D magnetization-prepared rapid gradient-echo (MPRAGE) T1 anatomical sequence (repetition time (TR)/time to echo (TE)/inversion time (TI) = 3000/3.7/905 ms, flip angle = 9°, voxel size = 1 mm isotropic, field of view (FOV) = 256 × 224 × 180 mm), a fluid-attenuated inversion recovery (FLAIR) sequence (TR/TE/TI = 9000/90/2500 ms, flip angle = 90°, voxel size = 0.94 × 0.94 mm FOV = 240 × 191 × 144 mm, slice thickness = 3mm), and a combined T2-weighted and proton density (PD) scan (TR/TE1/TE2 = 2500/9.5/90 ms, flip angle = 90°, voxel size = 0.94 × 0.94 mm, FOV = 240 × 191 × 144mm, slice thickness = 3mm). The following quantitative imaging sequences were acquired: high-angular resolution diffusion imaging (HARDI) sequence across 60 non-collinear diffusion gradients (b-value = 700 s/mm^2^, TR/TE = 7094/60ms, voxel size = 2mm isotropic, FOV = 224 × 224 × 154mm, slice thickness = 2.2mm), and a 32-echo gradient or 48-echo gradient- and spin-echo (GRASE) sequence (32-echo: TR/TE = 1000/10ms, reconstructed voxel size = 1 × 1 × 2.5mm, FOV = 230 × 190 × 100mm; 48-echo: TR/TE = 1073/8ms, reconstructed voxel size = 1 × 2 × 2.5mm, FOV = 230 × 190 × 100mm). Preliminary analyses of our data indicated there was no change in MWF or GMT2 estimates between the 32 and 48 echo GRASE sequences, which is consistent with previous reliability work done by our imaging group^27^; however, the scanner type may impact data due to difference in signal to noise ratio from different coil channel numbers, therefore we controlled for scanner type in all analyses.

### MRI preprocessing

Structural segmentation was performed with a previously published and validated segmentation and parcellation pipeline^28–30^. Briefly, T1, FLAIR, T2 and PD scans were linearly co-registered and supratentorial cerebral tissue was segmented into cerebrospinal fluid (CSF; sulcal and ventricular), grey matter, NAWM, and WMHs. WMHs were sub-classified as periventricular WMHs when the WMH made contact with ventricular CSF in three-dimensional space, and deep WMHs when the WMH did not make contact with ventricular CSF^29^. Stroke lesions were manually delineated over co-registered T1 and FLAIR scans by a single experienced researcher (by J.K.F).

Diffusion tensor images were preprocessed with FMRIB’s Software Library (FSL) diffusion toolbox (FDT)^31^. Briefly, DTI data were corrected for motion and eddy-current distortion, and the unweighted volume was skull-stripped using the Brain Extraction Toolbox (BET). FA and MD maps were generated using DTIFIT. T1 scans were co-registered with the DTI volume with no diffusion weighting (b=0 volume) by a rigid-body linear registration with a correlation ratio cost function using FSL’s FLIRT^32^. Registration quality was visually checked by a single rater and registrations were manually adjusted where necessary using tkregister^33^ (Freesurfer v.6.0, http://surfer.nmr.mgh.harvard.edu/).

Myelin water imaging was performed by fitting T2 relaxation distributions using a regularized nonnegative least-squares algorithm^34^ and extended phase graph algorithm^35^ with in-house MATLAB software that can be requested from https://mriresearch.med.ubc.ca/news-projects/myelin-water-fraction/. MWF was defined as the area under the T2 distributions from 10-40ms, relative to the total area of the T2 distribution. Intra/extracellular water geometric mean T2 (GMT2) was defined as the amplitude-weighted mean on a logarithmic scale of the T2 distribution between 40-200ms. T1 scans were co-registered with the first echo from the myelin water imaging scan, using a rigid-body linear registration with a mutual information cost function using FSL’s FLIRT^32^. Registration quality was visually checked by a single rater (by J.K.F).

### Regions of interest

See Figure 1 for an overview of the MRI processing pipeline and region of interest (ROI) creation. The objective of our ROI analysis was to examine whether a spatial gradient of changes to DTI and myelin water imaging metrics exists in WMHs and perilesional NAWM, as a function of distance from the WMH. We created five dilated NAWM masks in 2mm increments, from 2mm to 10mm of distance from the WMH. Smaller masks were subtracted from larger masks to create rings of tissue specific to each 2mm segment. We then used these dilated masks to subdivide the NAWM segmentation, creating ROIs of rings of increasing distance in 2mm increments in NAWM only^16^. To minimize partial volume effects from CSF contamination, CSF masks were dilated by 1mm and subtracted from tissue segmentations prior to analysis. To minimize the impact of the stroke lesion, stroke lesion masks were dilated by 10mm and removed from tissue segmentation prior to analysis. ROIs were then linearly registered to DTI and myelin water imaging space using the registration procedures detailed in the “MRI preprocessing” section. White matter metrics of interests (DTI: FA, MD; Myelin water imaging: MWF, GMT2) were extracted in DTI and myelin water imaging space, respectively.

**Figure 1:**
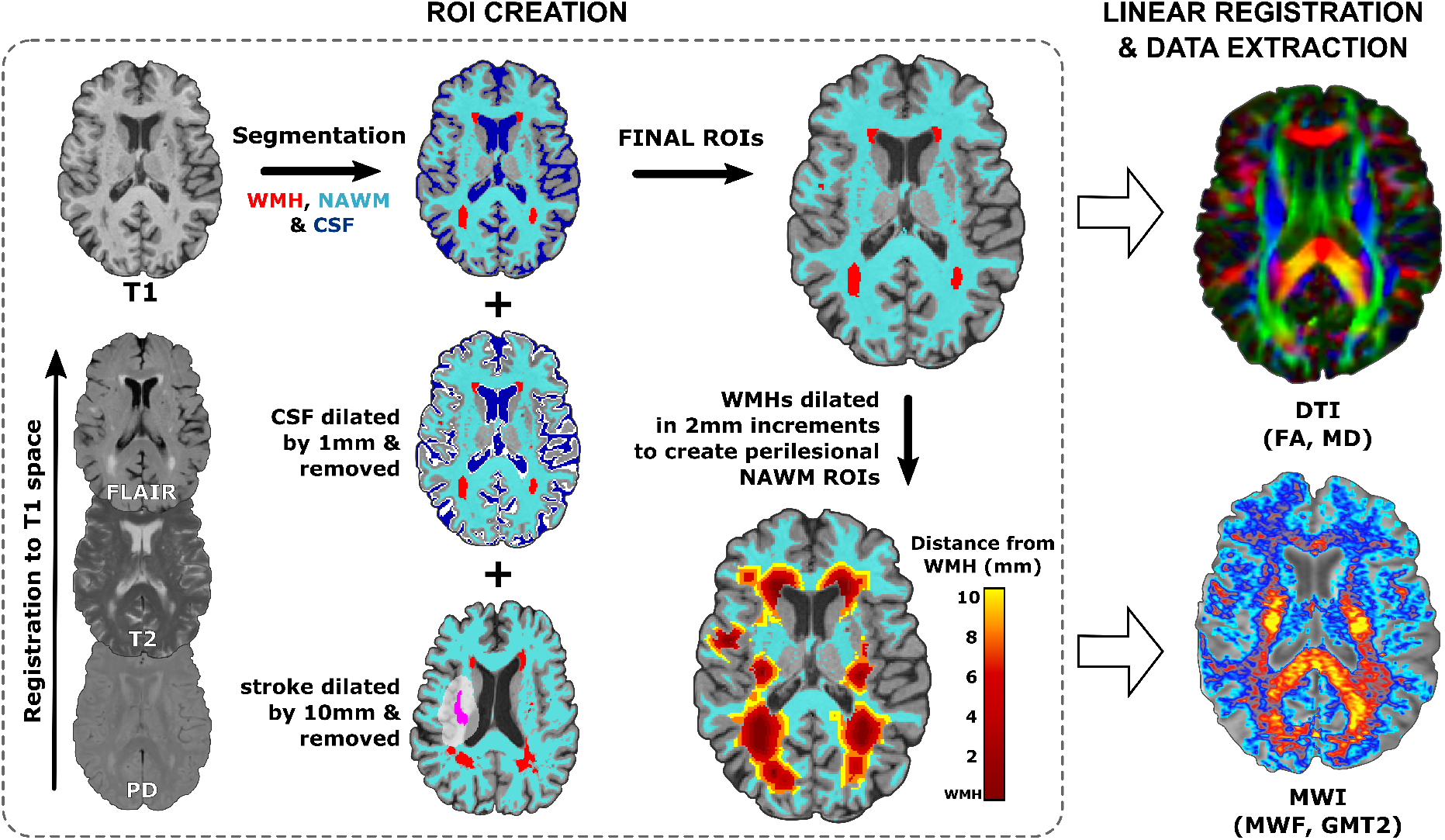
Overview of MRI processing pipeline and ROI creation. Tissue segmentation was performed in T1 space, CSF masks were dilated by 1mm and removed from tissue segmentations. For individuals in the chronic stroke group, stroke lesion masks were dilated by 10mm and removed from tissue segmentations. Next, WMH segmentations were serially dilated in 2mm increments from 2-10mm and extracted from NAWM to perform a spatial analysis of white matter metrics in perilesional NAWM as a function of distance from the WMH. ROIs were then linearly registered to DTI and MWI space, and mean data were extracted from ROIs in DTI and MWI space, respectively

### Statistical analysis

Statistical analyses were performed with R (programming environment v4.0.4). All data were checked for normality and whole-brain WMH volumes were log transformed prior to analysis. Predictor variables were group mean centered and standardized prior to analysis. We tested our hypotheses with a series of linear mixed effects models; the fixed effects of interest varied for each model and are described in subsequent sections. Separate models were performed with each white matter metric as an outcome measure (FA, MD, MWF & GMT2). All models had age and MoCA score entered as fixed effects. MRI scanner was entered as a random effect, which allowed us to combine data collected across two scanners while accounting for any potential effects of scanner. For models with repeated measures, participant was entered as a nested random effect within the random effect of MRI scanner. Linear mixed effects models were fit with the R packages lme4^36^ and lmerTest^37^, and the significance of predictors in the model were assessed with Satterthwaite’s approximation^38^. Posthoc contrasts for linear mixed effects models were performed with Tukey’s PSD adjustments using the R multicomp package^39^. The alpha threshold for significance was set at *p* < 0.05.

#### Spatial gradient of white matter metrics in WMHs and perilesional NAWM

We tested if white matter metrics varied by distance from the WMH (in WMH and perilesional NAWM) or by group (older adults vs chronic stroke). The factors Distance (WMH, 2mm, 4mm, 6mm, 8mm, 10mm) × Group (older adult vs chronic stroke) were entered as fixed effects. Distance was fit as a quadratic contrast, as we expected to see a non-linear change in white matter metrics with increasing distance^15,16^. Significant main effects of Distance were interrogated with posthoc contrasts comparing mean white matter metrics between each Distance level. Significant Distance × Group interactions were interrogated with Tukey posthoc contrasts comparing mean white matter metrics between each level of Distance within each Group, and between each Group within each level of Distance.

To test if WMH-related effects in individuals with chronic stroke were driven by the presence of a stroke lesion, we performed a secondary analysis of white matter metrics split by cerebral hemisphere (ipsilesional vs contralesional) limited to participants with unilateral stroke lesions. The factors Distance (WMH, 2mm, 4mm, 6mm, 8mm, 10mm; quadratic contrast) × Hemisphere (ipsilesional vs contralesional) were entered as fixed effects of interest. Planned posthoc contrasts for significant Distance × Group interactions compared mean white matter metrics between each level of Distance within each Hemisphere, and between each Hemisphere within each level of Distance.

#### Relationships between WMH volume and white matter metrics

Finally, we tested whether white matter metrics in the WMH relate to the severity of white matter disease, indexed by whole-brain supratentorial WMH volume. This analysis was performed across the entire sample, on data from the WMH ROI. We ran linear mixed effects models with WMH volume (log transformed) entered as the fixed effect of interest. As this model did not contain repeated measures across subjects, only scanner was included as a random effect.

### Data availability

The data supporting the findings of this study are available on request from the corresponding author. The data are not publicly available because they contain information that could compromise the privacy of research participants.

## Results

### Participants

50 older adults (ages 51-80) and 33 individuals with chronic stroke (ages 45-80) participated in this study. 54 participants were scanned on the Phillips Achieva scanner, and 26 were scanned on the Phillips Elition scanner. Two older adults were excluded because their WMH volumes exceeded our ≥10 mL cut-off, and one older adult was excluded because they did not have any WMHs voxels identified in WMH segmentation, leaving a final sample size of 47 older adults. All participants were successfully processed through MRI pre-processing steps.

Table 1 presents participant demographics. Relative to older adults, individuals with chronic stroke had a lower percentage of females in the sample, lower cognitive function (indexed by MoCA scores), and larger WMH volumes. Supplementary Figure 1 presents lesion overlap images for WMHs and stroke lesions across the sample. Supplementary Table 1 presents tests for within-group sex differences in white matter metrics. There were no significant sex differences in whole brain WMH or stroke volumes or in white matter metrics in the WMH in either group (all *p* > 0.05).

**Table 1:** Participant demographics. P-values presented are results of independent sample t-tests for age, MoCA & WMH volume, and chi-squared test for Gender. Age is in years and time since stroke is in months. Bold values indicate statistical significance (*p* < 0.05)

|  | Older adult group | Chronic stroke group | *p* |
| --- | --- | --- | --- |
| n | 47 | 33 |  |
| Age<br>mean (SD) | 65 (7) | 67 (8) | 0.105 |
| Sex<br>n (%) females | 30 (64%) | 11 (33%) | **0.014** |
| MoCA<br>mean (SD)<br>% $\geq 26$ | 27 (2)<br>78% | 25 (3)<br>61% | **0.005** |
| WMH, mL<br>median (IQR) | 0.352<br>[0.124- 0.599] | 2.676<br>[1.411-10.444] | **< 0.001** |
| Time since stroke<br>mean (SD) | n/a | 76 (57) |  |

### There is a spatial gradient of fluid-sensitive white matter metrics in WMHs and perilesional NAWM

Figure 2.A presents mean data for DTI and myelin water imaging white matter metrics (DTI: FA & MD; myelin water imaging: MWF & GMT2) across WMHs and perilesional NAWM (2-10mm segments) between older adults and individuals with chronic stroke. Table 2 presents results from linear mixed effects models assessing the effect of Distance (from the WMH), Group (older adult vs chronic stroke), and Distance^*^Group interactions on each white matter metric.

**Table 2:**
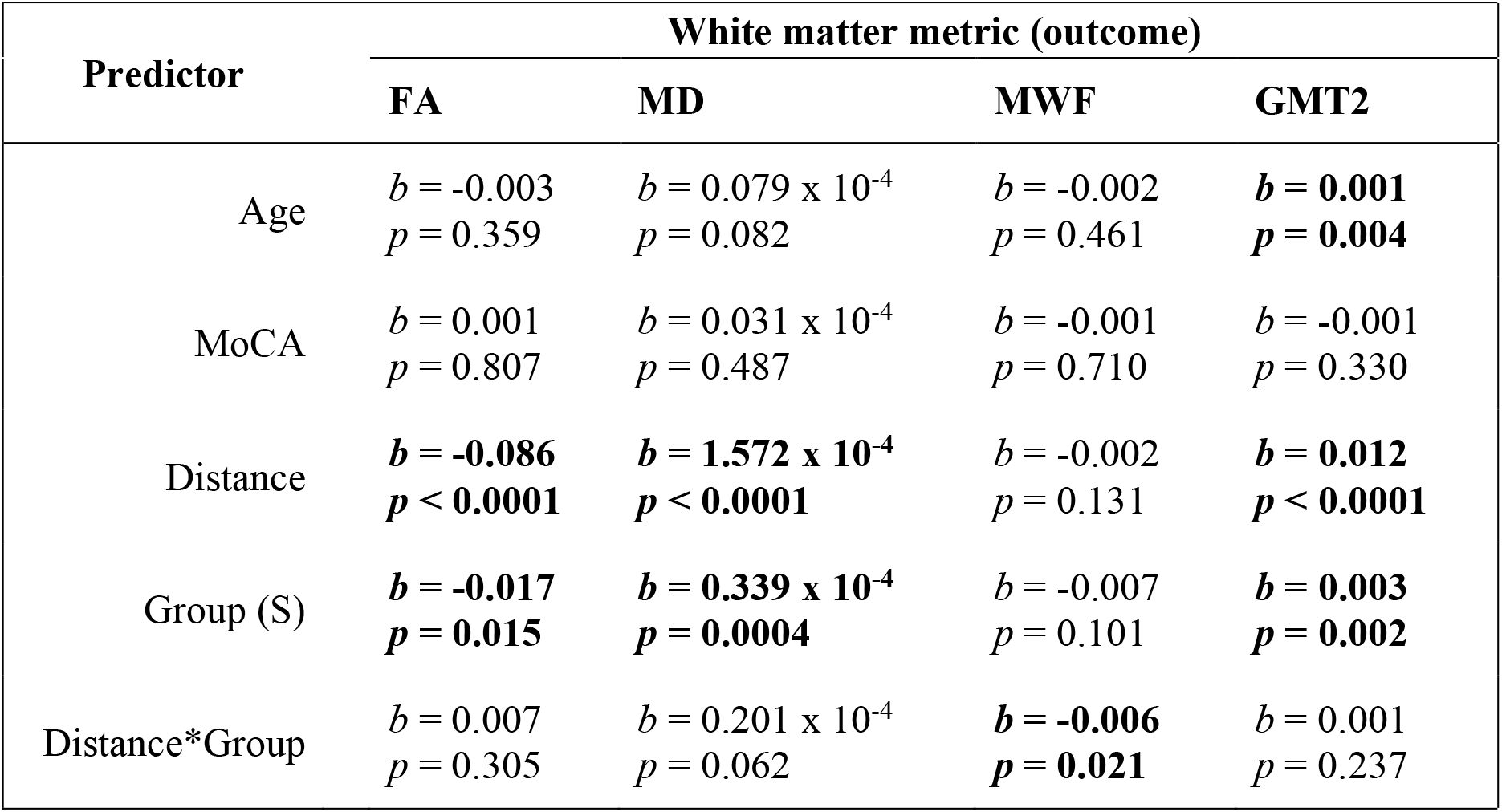
Distance × Group models. Results from linear mixed effects models testing relationships between white matter metrics in the WMH and perilesional NAWM, as a function of Distance (in 2mm increments) from the WMH, between Groups (older adults and chronic stroke, with stroke group (S) as the reference category). Cells present standardized parameter estimates and p-values. Bold values indicate statistical significance (*p* < 0.05)

**Figure 2:**
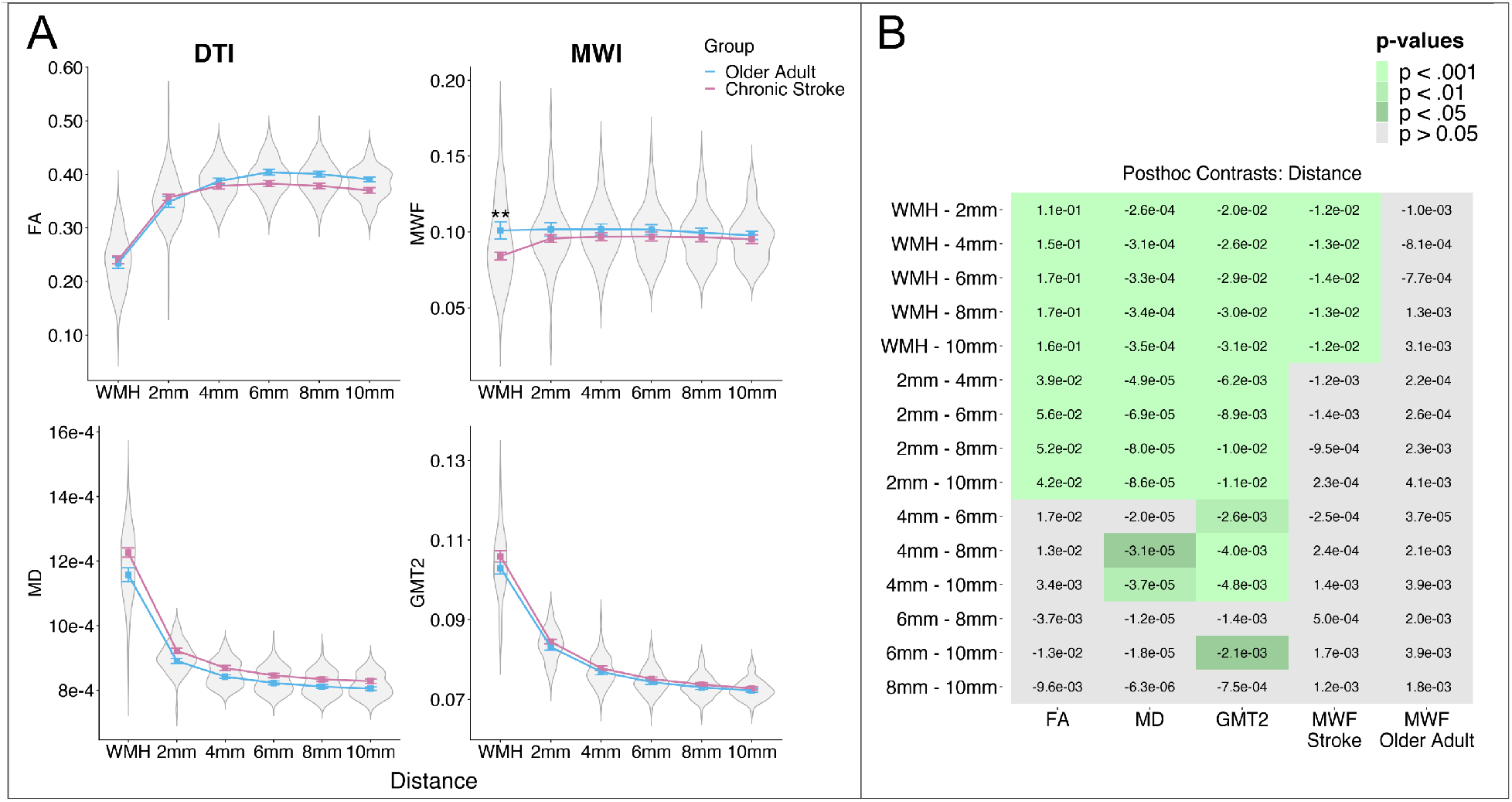
Spatial gradient of white matter metrics in WMHs and perilesional NAWM. **(A)** Mean DTI (left) and myelin water imaging (right) metrics plotted as a function of Distance from the WMH by Group (older adults vs. chronic stroke). FA, MD and GMT2 changed as a function of distance from the WMH (higher FA, lower MD and GMT2 with increasing Distance from the WMH). MWF in the WMH was significant lower in individuals with chronic stroke relative to older adults. Asterisks represent significant posthoc contrast for Group × Distance interactions (^**^= p < 0.010, see Table 2). **(B)** Posthoc contrasts comparing mean white matter metrics for main effects of Distance (FA, MD & GMT2) and Distance separated by Group effects for outcome metrics with significant Distance^*^Group interactions (MWF). Cell values are the linear estimates for each contrast, and green cells indicate significant contrasts.

There was a main effect of Distance on white matter metrics for FA, MD and GMT2. FA was lowest in the WMH and showed a quadratic increase in perilesional NAWM as a function of distance from the WMH. MD and GMT2 were highest in the WMH, with a quadratic decrease in perilesional NAWM as a function of distance from the WMH (Table 2; Figure 2.A). FA, MD and GMT2 also showed a main effect of Group, with lower FA and higher MD and GMT2 in the chronic stroke group relative to the older adult group (Table 2; Figure 2.A), with no significant Distance^*^Group interaction effects (all *p* > 0.05, Table 2). Tukey adjusted posthoc contrasts are visualized in Figure 2.B. Posthoc contrasts for the main effect of Distance revealed that FA MD, and GMT2 values were significantly different in the WMH relative to all 5 NAWM segments, and in the 2mm NAWM segment relative to all remaining NAWM segments (4-10mm; Figure 2.B). Additionally, GMT2 in the 4mm NAWM segment was significantly higher than GMT2 in all remaining NAWM segments (6-10mm; Figure 2.B).

MWF did not show a main effect of Distance, but instead showed a significant Group^*^Distance interaction (Table 2). Posthoc contrasts revealed that within the WMH ROI there was significantly lower MWF in the chronic stroke group relative to the older adult group (*t*_45_ = 3.284, *p* = 0.002; Figure 2.A). Additionally, within the chronic stroke group MWF in the WMH was significantly lower than in all perilesional NAWM segments, whereas within the older adult group there were no significant differences in MWF values between the WMH and perilesional NAWM (Figure 2.B).

### The spatial gradient is not driven by CSF partial volume effects from proximity to the cerebral ventricles

We confirmed that our results for fluid-sensitive metrics (FA, MD and GMT2) were not driven by fluid (i.e., CSF) contamination from partial volume effects in WMHs bordering the cerebral ventricles. We conducted a supplementary analysis of deep WMHs only, which are WMHs that do not contact the cerebral ventricles in three-dimensional space. We restricted this analysis to the older adult group, because in individuals with chronic stroke deep WMHs may be associated with stroke lesions which could confound these results.

Results of linear mixed effects models within deep WMHs testing for the effects of Distance on white matter metrics are presented in Supplementary Figure 2 and Supplementary Table 2 and briefly summarized here. Results from the deep WMH analysis were consistent with the results of the primary analysis across all WMHs, such that FA, MD and GMT2 showed a spatial effect of Distance on white matter metrics, whereas MWF showed no significant effect of Distance (see: Supplementary Figure 2 and Table 2).

### The spatial gradient does not differ by cerebral hemisphere in the chronic stroke group

To confirm that results for individuals with chronic stroke were not driven by the presence of stroke lesions, we divided the chronic stroke group data by hemisphere (ipsilesional and contralesional) and performed a secondary analysis examining the impact of Distance and Hemisphere on white matter metrics in WMHs and perilesional NAWM limited to individuals with unilateral stroke. Eleven participants in our sample had bilateral lesions and were excluded, leaving a sample size of 22 for this analysis.

Figure 3 presents mean data from white matter metrics (FA, MD, MWF & GMT2) across WMHs and perilesional NAWM (2-10mm segments) between the contralesional and ipsilesional hemisphere for individuals with unilateral stroke lesions. Table 3 presents results from linear mixed effects models assessing the effect of Distance (from the WMH), Hemisphere (ipsilesional vs contralesional), and Distance^*^Hemisphere interactions on white matter metrics.

**Table 3:** Distance × Hemisphere models in individuals with unilateral stroke. Results from linear mixed effects models conducted for individuals with unilateral chronic stroke lesions (n = 22) testing relationships between white matter metrics in the WMH and perilesional NAWM, as a function of Distance from the WMH, between Hemispheres (ipsilesional and contralesional, with ipsilesional (I) as the reference category). Cells present standardized parameter estimates and p-values. Bold values indicate statistical significance (*p* < 0.05).

| Predictor | White matter metric (outcome) |  |  |  |
| --- | --- | --- | --- | --- |
|  | FA | MD | MWF | GMT2 |
| Age | $b = -0.012$<br>$p = 0.054$ | $b = 0.087 \times 10^{-4}$<br>$p = 0.390$ | $b = -0.001$<br>$p = 0.733$ | $b = \mathbf{0.002}$<br>$p = \mathbf{0.008}$ |
| MoCA | $b = -0.007$<br>$p = 0.421$ | $b = 0.131 \times 10^{-4}$<br>$p = 0.340$ | $b = -0.004$<br>$p = 0.541$ | $b = -0.001$<br>$p = 0.672$ |
| Distance | $b = \mathbf{-0.078}$<br>$p < \mathbf{0.0001}$ | $b = \mathbf{1.810 \times 10^{-4}}$<br>$p < \mathbf{0.0001}$ | $b = \mathbf{-0.008}$<br>$p = \mathbf{0.0003}$ | $b = \mathbf{0.013}$<br>$p < \mathbf{0.0001}$ |
| Hemisphere (I) | $b = 0.002$<br>$p = 0.544$ | $b = 0.057 \times 10^{-4}$<br>$p = 0.323$ | $b = 0.001$<br>$p = 0.578$ | $b = -0.001$<br>$p = 0.322$ |
| Distance*Hemisphere | $b = -0.005$<br>$p = 0.619$ | $b = -0.049 \times 10^{-4}$<br>$p = 0.729$ | $b = 0.001$<br>$p = 0.679$ | $b = -0.001$<br>$p = 0.137$ |

**Figure 3:**
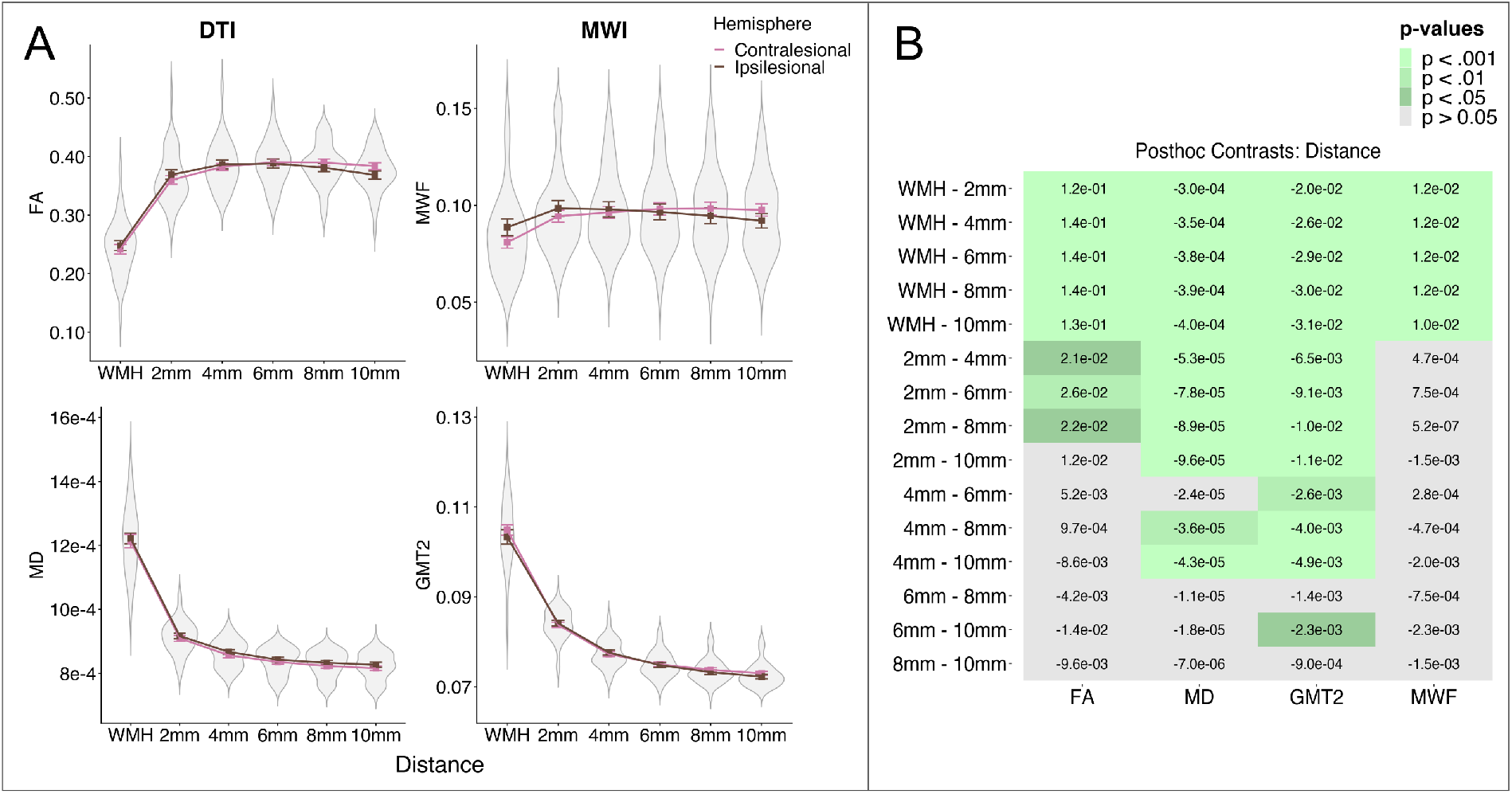
Spatial gradients of white matter metrics between the cerebral hemispheres for individuals with unilateral stroke. **(A)** Mean DTI (left) and myelin water imaging (right) metrics for individuals with unilateral chronic stroke lesions (*n* = 22). Data are plotted as a function of Distance from the WMH by Hemisphere (ipsilesional vs. contralesional). All white matter metrics changed as a function of distance from the WMH (higher FA and MWF and lower MD and GMT2 with increasing Distance from the WMH; see Table 3). The spatial gradient in WMH and perilesional NAWM does not differ in the ipsilesional hemisphere, and thus was not driven by the presence of stroke lesions. **(B)** Posthoc contrasts comparing mean white matter metrics for main effects of Distance. Cell values are the linear estimates for each contrast, and green cells indicate significant contrasts.

Results from this sub-analysis were consistent with the results from our full sample analysis. There was a main effect of Distance on all white matter metrics (FA, MD, MWF, LT2), with no main effect of Hemisphere or Hemisphere^*^Distance interactions (Table 3; Figure 3). The spatial gradients of white matter metrics in WMH and perilesional NAWM were not different between the cerebral hemispheres in individuals with unilateral chronic stroke.

### Relationships between WMH volume and white matter metrics

Figure 4 presents relationships between white matter metrics (FA, MD, MWF & GMT2) in the WMH and whole brain WMH volume. Results of linear mixed effects analyses assessing the relationship between white matter metrics in the WMH and whole-brain WMH volume are presented in Table 4. FA did not relate to WMH volume. MD and GMT2 had a significant positive relationship with WMH volume, such that higher WMH volume was related to higher MD and GMT2 in the WMH (Figure 4, Table 4). MWF had a significant negative relationship with WMH volume, such that higher WMH volume was associated with lower MWF within the WMH (Figure 4, Table 4).

**Table 4:** Relationships between WMH lesion metrics and whole-brain WMH volume. Results from linear mixed effects models testing relationships between white matter metrics in the WMH and whole-brain WMH volume (log transformed). Cells present standardized parameter estimates and p-values. Bold values indicate statistical significance (*p* < 0.05)

| Predictor | White matter metric (outcome) |  |  |  |
| --- | --- | --- | --- | --- |
|  | FA | MD | MWF | GMT2 |
| Age | $b = 0.001$<br>$p = 0.985$ | $b = -0.074 \times 10^{-4}$<br>$p = 0.585$ | $b = 0.002$<br>$p = 0.406$ | $b = 0.002$<br>$p = 0.074$ |
| MoCA | $b = -0.001$<br>$p = 0.913$ | $b = 0.025 \times 10^{-4}$<br>$p = 0.843$ | $b = -0.002$<br>$p = 0.536$ | $b = -0.001$<br>$p = 0.217$ |
| WMH volume | $b = 0.006$<br>$p = 0.445$ | $b = \mathbf{0.591 \times 10^{-4}}$<br>$p < \mathbf{0.001}$ | $b = \mathbf{-0.007}$<br>$p = \mathbf{0.031}$ | $b = \mathbf{0.004}$<br>$p < \mathbf{0.001}$ |

**Figure 4:**
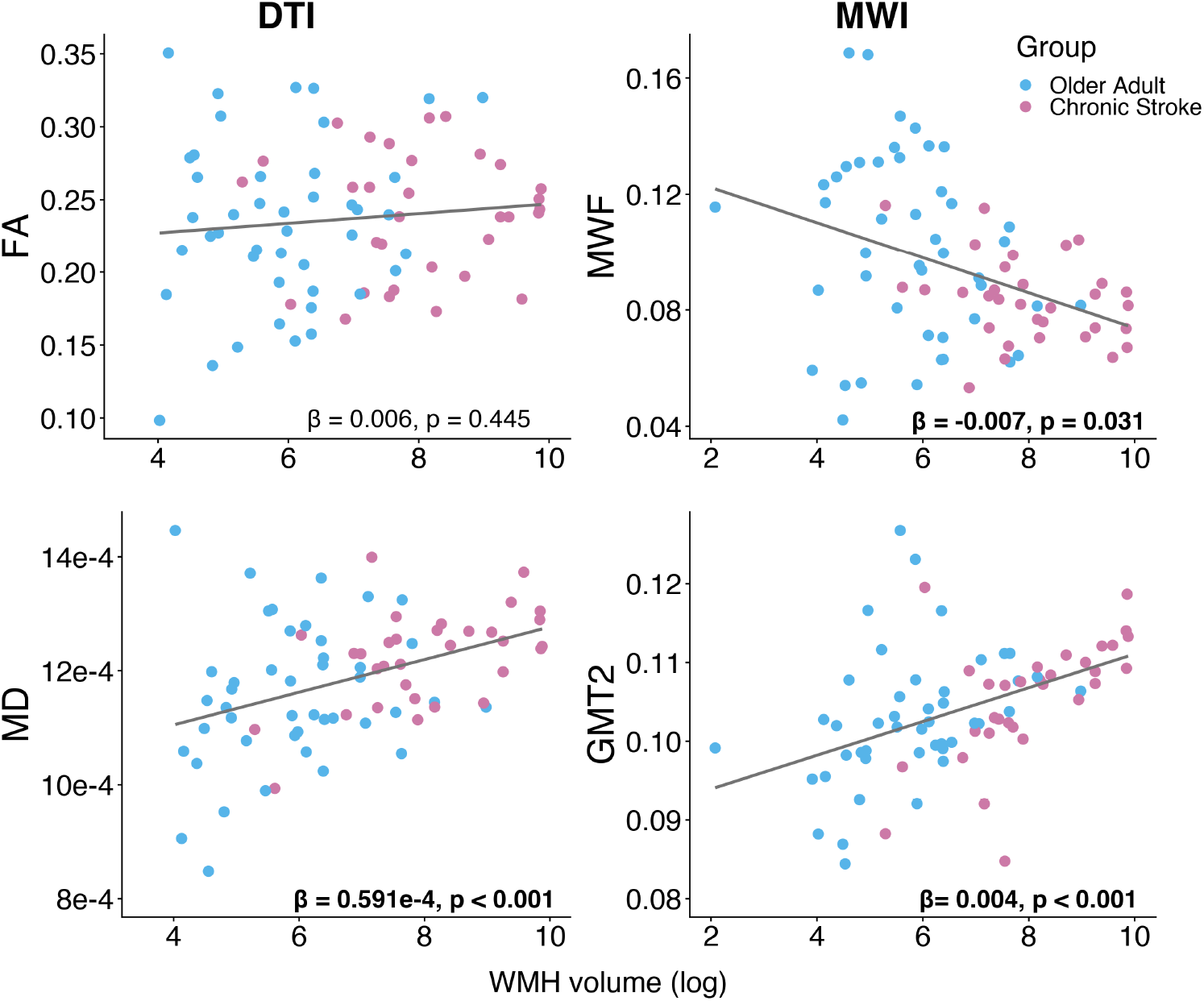
Relationships between white matter metrics within the WMH lesion and whole-brain WMH volume. Blue points are older adults and pink points are individuals with chronic stroke. The text presents parameter estimates and p-values for WMH volume (log transformed) from the associated linear mixed effects models analysis (see Table 4), bolded text indicates significant relationships.

## Discussion

In the current study we combined DTI and myelin water imaging to measure subvisible changes to tissue characteristics in WMHs and perilesional NAWM. Our study has two important findings. First, we confirm the existence of a so-called “WMH penumbra”^15^ in DTI metrics in the perilesional NAWM^15,16,40,41^. We observed with myelin water imaging that these penumbral changes in perilesional NAWM appear only for measures of interstitial fluid (GMT2), not demyelination (MWF), including in more severe stages of cerebrovascular disease (individuals with chronic stroke). Second, we found that demyelination does occur in the WMH itself, but only in individuals with more severe stages of cerebrovascular disease; both within the chronic stroke group and observed as a continuous relationship with whole brain WMH volume across the entire sample.

### Spatial effects of WMHs in perilesional white matter

Here we show that altered white matter microstructure in perilesional NAWM reflects increased interstitial fluid, and not demyelination. Consistent with previous work^15,16^, FA and MD are altered in perilesional NAWM. Myelin water imaging further revealed a strong spatial gradient of GMT2, which paralleled MD levels in the direction and amplitude of effects. This suggests that interstitial fluid accumulation extends beyond the WMH lesion into perilesional NAWM. GMT2 was significantly elevated in NAWM up to 4mm from the WMH. By contrast, DTI metrics were altered only at 2mm distance from the WMH. Importantly, we observed these spatial effects in both groups, including in our cohort of older adult participants with mild WMHs volumes. WMHs typically show expansion over time, though the rate of expansion is variable, and recent data suggests WMH lesions can also remain stable or even regress over a 1-5 year period^42^. FA is lower in perilesional NAWM that goes on to become a WMH, compared to NAWM that remains stable at follow-up^41^. Our findings suggest that GMT2 may have similar, or greater, sensitivity than FA to detecting interstitial fluid changes in perilesional NAWM, which may predict individual profiles of WMH progression over time. Further, our finding of elevated GMT2 in perilesional NAWM supports theories that early WMHs are potentially reversible with resolution of interstitial edema^8,43^. Thus, fluid-sensitive MRI markers may be useful as early indicators of future WMH expansion. This has potential for development as a screening tool for early WMH formation in individuals who do not yet show clinically meaningful amounts of WMH.

While the spatial effects in WMH and perilesional NAWM were observed for fluid-sensitive metrics (FA, MD & GMT2), fluid contamination from nearby ventricular CSF was not responsible for these results. Our supplementary analysis confirmed that an identical spatial gradient of change was observed in perilesional NAWM surrounding deep WMHs, which are randomly distributed throughout the cerebral grey matter, and are not adjacent to the cerebral ventricles. Therefore, our data strongly suggest that WMHs are characterized by local fluid accumulation, which extends in a penumbra-like fashion into neighboring NAWM.

MWF, in contrast with the fluid-sensitive metrics, did not show a spatial gradient of effects in perilesional NAWM. Rather, lower MWF was only observed in the WMH lesion of individuals with more severe cerebrovascular disease (individuals with chronic stroke), and in these individuals the low MWF levels did not extend to perilesional NAWM. This finding suggests that overt demyelination and neuronal damage occur only in more advanced WMH disease stages, and early WMH-related pathology in the NAWM is not related to demyelination.

Our findings in perilesional NAWM are congruent with a theory of WMH formation stemming from vascular stiffening and increased blood brain barrier permeability, leading to leakage of fluid into interstitial spaces^44^. Histopathological evidence suggests that WMH formation is driven by pathology to the blood vessels, including vessel tortuosity and venous collagenosis^9,10,45,46^. In a recent MRI study, Wong *et al*^47^ reported increased blood brain barrier leakage and reduced cerebral blood flow in perilesional NAWM, with a similar spatial gradient of effects with increasing distance from the WMH that we observed in MD, FA and GMT2 data. In summary, our findings are supported by theories of WMH formation from the histopathological literature and contribute to a body of evidence showing penumbral effects of fluid accumulation in perilesional NAWM surrounding WMHs.

### White matter markers in the WMH lesion

We found that within the WMH there are subvisible changes in tissue characteristics reflecting both increased interstitial fluid and demyelination. With increasing WMH volume (i.e., increasing severity of WMHs) there is increased interstitial fluid (MD & GMT2) and decreased myelination (MWF) in the WMH lesion. Previous histological work has shown varying degrees of demyelination within the WMH that does not reliably relate to the severity of hyperintensity on a T2-weighted scan^10,48^. This underscores why T2-weighted imaging alone is insufficient for staging WMH progression. FA in the WMH also does not relate to histologically measured myelin concentration^40^. A specific neuroimaging myelin marker such as MWF might be able to inform about evolving pathology in the WMH as demyelination progresses. MWF may be a prospective measure of WMH severity, and future studies should test whether MWF in the WMH is a better predictor of functional decline than whole-brain WMH volumes.

### Implications for quantitative neuroimaging research of WMHs

Given the low specificity of DTI markers, our findings highlight that the biological significance of altered FA and MD must always be interpreted with caution. Despite this caveat, FA and MD are sensitive metrics that can inform about WMH progression. Longitudinal studies have shown that NAWM tissue which converts to WMH at follow-up shows reduced FA and higher MD at baseline, relative to NAWM that does not contain WMH at follow-up^41,49,50^. Thus, DTI can detect changes to tissue microstructure *before* a WMH appears on a T2-weighted scan. DTI is attractive tool for clinical translation because it is commonly employed, and diffusion weighted imaging is already used as a diagnostic tool in clinical settings. The challenge for the field will be to delineate how and where DTI might be applied to maximize early detection of WMHs or in tracking response to therapeutic interventions before irreversible myelin loss has occurred.

To our knowledge, we are the first study to test differences between MWF levels in WMHs of presumed vascular origin and NAWM. Myelin water imaging is a novel imaging technique, and only a handful of previous studies have applied myelin water imaging in typical aging and dementia. Kavroulakis *et al*^51^ found MWF in NAWM is reduced in individuals with Alzheimer’s Disease (without significant WMHs), relative to cognitively intact older adults. Similarly, Dao *et al*.^52^ found that MWF in NAWM relates to processing speed in older adults with WMHs. These studies suggest that MWF levels in broad white matter networks may be important in maintaining cognitive performance. In light of the current findings, we suggest that MWF levels in the WMH might also be useful as an *in vivo* marker of demyelination for WMH disease staging.

### Limitations

Our study has several limitations. First, the cohort of individuals with chronic stroke was included to examine how the severity of cerebrovascular disease may impact white matter markers in WMHs, yet it is possible that overt stroke lesions could confound results. We took several steps to confirm that our results were not driven by the presence of overt stroke lesions. We dilated each stroke lesion mask by 10mm and removed this tissue from analysis, to avoid measuring from tissue in the perilesional stroke infarct zone. Further, we split our ROIs by hemisphere (ipsilesional vs. contralesional) and results were consistent within each hemisphere. We also observed (Supplementary Figure 1) that WMHs in the individuals with chronic stroke were symmetrical and resembled the typical topography of WMH observed in more severe stages of WMHs in typically-aging adults^53^. Therefore, we believe our findings will generalize to severe WMH non-stroke populations, which should be confirmed in future research. Second, our study employed *in* vivo neuroimaging, and we cannot confirm the present findings with more objective, albeit invasive, histological techniques. However, MWF has previously been histologically validated as a sensitive marker of myelin content^21,22^, so we have high confidence in the validity of interpretation of our myelin water fraction findings for fluid and myelin content in the tissue. Finally, combining data across multiple scanners may be considered a limitation in the introduction of noise into the data. However, this approach also has strengths as it allowed us to maximize power and reduce research waste, and any variability associated with scanners was accounted for in our statistical models.

## Conclusions

Here we show that quantitative neuroimaging can be used to investigate underlying pathology in WMHs and perilesional NAWM. We observe that the NAWM bordering the WMH shows tissue characteristics that suggests a transitionary or intermediate stage between healthy NAWM tissue and WMH lesions. We found that this intermediate stage is characterized by increased interstitial fluid, and not decreased myelin concentration. Within the WMH lesion tissue characteristics vary depending on the severity of cerebrovascular disease, and myelin concentration is reduced in the WMH lesion only in individuals with more severe cerebrovascular disease. Our findings have two potential clinical applications. First, fluid-sensitive metrics (FA, MD and/or GMT2) could be used to identify NAWM at risk of future transition to a WMH lesion. Second, MWF levels in the WMH lesion capture variability in the severity of WMH pathology (i.e., the degree of demyelination) which could provide a method to measure the severity of WMH beyond whole-brain lesion volumes.

## Abbreviations

CSF: cerebrospinal fluid
DTI: Diffusion tensor imaging
FA: fractional anisotropy
GMT2: geometric mean T2
MD: mean diffusivity
MWF: myelin water fraction
NAWM: normal appearing white matter
ROI: region of interest
WMH: white matter hyperintensity

## Acknowledgements

We gratefully acknowledge the assistance of imaging analysts in the LC Campbell Cognitive Neurology Research Unit, including Sabrina Adamo, Miracle Ozzude, Dr. Fuqiang Gao, and Christopher Scott. We thank Dr. Teresa Liu Ambrose for helpful conversations related to this paper.

