## Supplementary material for "In vivo myelin imaging and tissue microstructure in white matter hyperintensities and perilesional white matter": Data Supplement

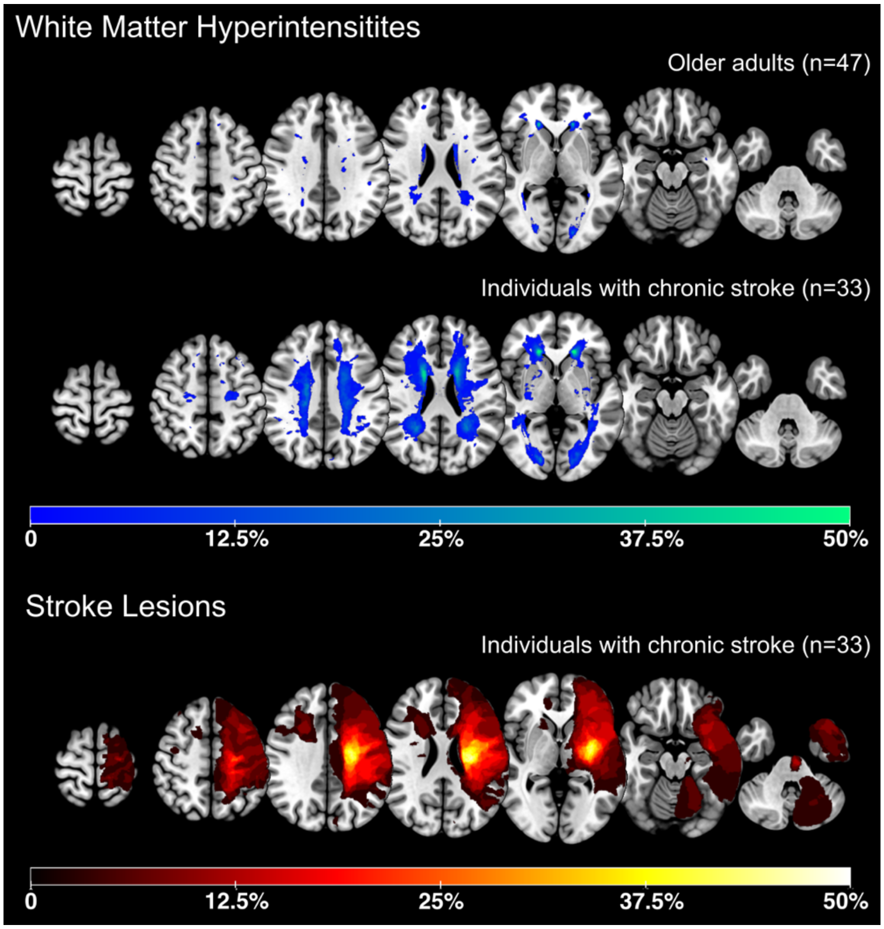


Supplementary Figure 1: Lesion overlap images for white matter hyperintensities (WMH; in blue) and stroke lesions (in red). WMH maps are split by group (older adults and individuals with chronic stroke). Stroke lesions were flipped along the L/R axis so that all symptomatic strokes (contralateral to the impaired upper extremity) were visualized in the left hemisphere (note: images are in radiological orientation). Color bar represents the percentage of the sample with a lesion in each voxel.

| | Older Adults | | | Individuals with Chronic Stroke | | |
|---|---|---|---|---|---|---|
| | Females (n = 30) | Males (n = 17) | *p* | Females (n = 11) | Males (n = 22) | *p* |
| WMH volume (mL) | 5.594 | 5.866 | 0.548 | 8.008 | 8.155 | 0.767 |
| Stroke volume (mL) | - | - | | 8.990 | 8.680 | 0.668 |
| FA | 0.223 | 0.255 | 0.076 | 0.250 | 0.235 | 0.339 |
| MD | 0.117 x $10^{-2}$ | 0.113 x $10^{-2}$ | 0.413 | 0.123 x $10^{-2}$ | 0.122 x $10^{-2}$ | 0.775 |
| GMT2 | 0.102 | 0.104 | 0.488 | 0.103 | 0.107 | 0.103 |
| MWF | 0.099 | 0.105 | 0.590 | 0.083 | 0.084 | 0.774 |

Supplementary Table 1: Sex differences in imaging metrics for older adults and individuals with chronic stroke, compared with independent samples t-tests.

### Is the spatial gradient in white matter changes driven by CSF partial volume effects due to proximity to the cerebral ventricles?

We performed this supplementary analysis to test whether results seen in WMH and perilesional NAWM were affected by partial volume concentration (i.e. CSF), due to the proximity of periventricular WMH to the cerebral ventricles. We restricted our analysis to the healthy older adult group, and tested data from deep WMHs only. Deep WMHs are WMHs that do not contact the cerebral ventricles in 3D space, are randomly distributed throughout cerebral tissue, and do not follow the stereotyped localization of WMHs adjacent to the cerebral ventricles.

34 healthy older adults had deep WMHs and were included in the supplementary analysis. Deep WMH masks were dilated by 2mm and used to segment NAWM, then linearly registered to DTI and myelin water imaging space, with identical methods as those employed for whole-brain WMHs. Detailed methods are provided in the body of the main document.

Supplementary Figure 2 presents mean data for DTI & myelin water imaging white matter metrics (DTI: FA & MD, myelin water imaging: MWF & GMT2) across WMHs and perilesional NAWM (2-10mm segments) for deep WMHs. Supplementary Table 1 presents results from linear mixed effects models assessing the effect of Distance (from the deep WMH) on white matter metrics. There was an effect of Distance on white matter metrics for FA, MD and GMT2, mirroring the findings of whole-brain WMHs. FA was lowest in the WMH and showed a quadratic increase in perilesional NAWM at greater distances from the WMH. MD and GMT2 were highest in the WMH, with a quadratic decrease in perilesional NAWM at greater distances from the WMH. There was no effect of distance on MWF concentration. (Supplementary Table 1; Supplementary Figure 2.A). Posthoc contrasts after Tukey's HSD adjustment for multiple comparisons are visualized in Supplementary Figure 2.B. Posthoc contrasts for the main effect of Distance revealed that FA MD, & GMT2 values were significantly different in the WMH relative to all 5 NAWM segments, and in the 2mm NAWM segment relative to all remaining NAWM segments (4-10mm; Supplementary Figure 2.B). For GMT2, GMT2 in the 4mm NAWM segment was significantly higher than GMT2 in all remaining NAWM segments (6-10mm; Supplementary Figure 2.B).

This pattern of findings is identical to the findings observed for the older adult group in our primary analysis, indicating that our primary results were not driven by CSF partial volume effects.

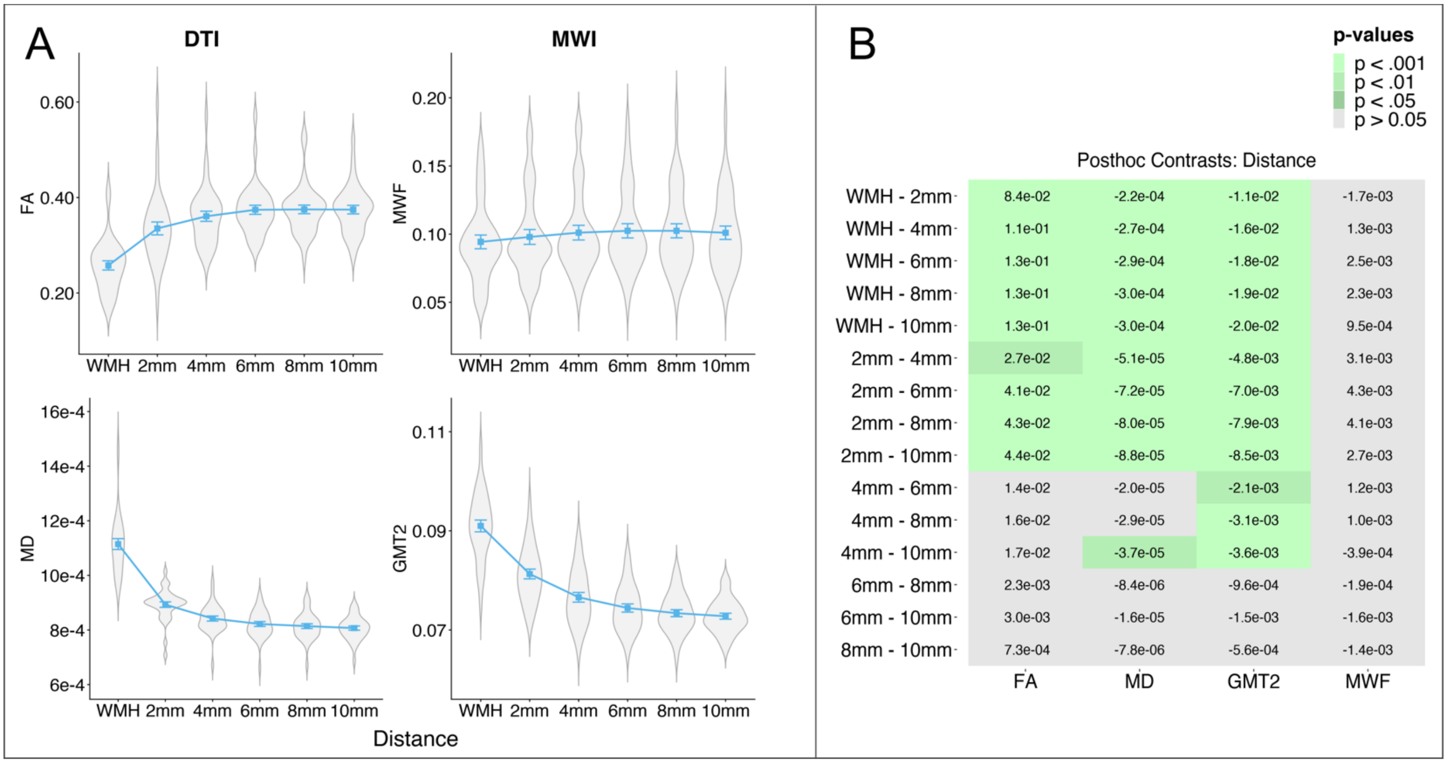


Supplementary Figure 2: A) Mean DTI (left) & myelin water imaging (right) white matter metrics plotted as a function of Distance from the deep WMH. FA, MD & GMT2 changed as a function of Distance from the deep WMH (higher FA and lower MD and GMT2 with increasing Distance from the WMH). B: Posthoc contrasts comparing mean white matter metrics between each level of Distance. Cell values are the linear estimates for each contrast, and green cells indicate significant contrasts.

| **Predictor** | **White matter metric (outcome)** | | | |
|---|---|---|---|---|
| | **FA** | **MD** | **MWF** | **GMT2** |
| Age | $b$ = -0.002<br>$p$ = 0.800 | $b$ = 0.102 x $10^{-4}$<br>$p$ = 0.110 | $b$ = 0.001<br>$p$ = 0.738 | $b$ = 0.114 x $10^{-3}$<br>$p$ = 0.886 |
| MoCA | $b$ = 0.009<br>$p$ = 0.233 | $b$ = 0.018 x $10^{-4}$<br>$p$ = 0.778 | ***b* = 0.010**<br>***p* = 0.021** | $b$ = 0.396 x $10^{-3}$<br>$p$ = 0.626 |
| Distance | ***b* = -0.056**<br>***p* < 0.0001** | ***b* = 1.324 x $10^{-4}$**<br>***p* < 0.0001** | $b$ = -0.001<br>$p$ = 0.627 | ***b* = 0.007**<br>***p* < 0.0001** |

Supplementary Table 2: Results from linear mixed effects models testing relationships between white matter metrics in deep WMHs and perilesional NAWM, as a function of Distance (in 2mm increments) from the deep WMH. Cells present standardized parameter estimates and p-values. Bold values indicate statistical significance ($p < 0.05$)
